# A standardized analytics pipeline for reliable and rapid development and validation of prediction models using observational health data

**DOI:** 10.1101/2021.03.23.21254098

**Authors:** Sara Khalid, Cynthia Yang, Clair Blacketer, Talita Duarte-Salles, Sergio Fernández-Bertolín, Chungsoo Kim, Rae Woong Park, Jimyung Park, Martijn Schuemie, Anthony Sena, Marc A. Suchard, Seng Chan You, Peter Rijnbeek, Jenna Reps

**Affiliations:** Centre for Statistics in Medicine. Nuffield Department of Orthopaedics, Rheumatology, and Musculoskeletal Sciences (NDORMS), University of Oxford, Oxford, UK. Botnar Research Centre; Erasmus University Medical Center; Observational Health Data Analytics, Janssen Research and Development, Titusville, NJ, USA; Fundació Institut Universitari per a la recerca a ľAtenció Primária de Salut Jordi Gol i Gurina; Fundació Institut Universitari per a la recerca a ľAtenció Primária; de Salut Jordi Gol i Gurina (IDIAPJGol), Barcelona, Spain; Department of Biomedical Sciences, Ajou University Graduate School of Medicine; 1. Observational Health Data Analytics, Janssen Research and Development, Titusville, NJ, USA 2. Department of Medical Informatics, Erasmus University Medical Center, Rotterdam, The Netherlands; University of California, Los Angeles; Department of Biomedical Informatics, Ajou University School of Medicine; Department of Medical Informatics, Erasmus University Medical Center, Rotterdam, The Netherlands

**Keywords:** COVID-19, data harmonization, data quality control, distributed data network, machine learning, risk prediction

## Abstract

**Background and Objective:** As a response to the ongoing COVID-19 pandemic, several prediction models have been rapidly developed, with the aim of providing evidence-based guidance. However, no COVID-19 prediction model in the existing literature has been found to be reliable. Models are commonly assessed to have a risk of bias, often due to insufficient reporting, use of non-representative data, and lack of large-scale external validation. In this paper, we present the Observational Health Data Sciences and Informatics (OHDSI) analytics pipeline for patient-level prediction as a standardized approach for rapid yet reliable development and validation of prediction models. We demonstrate how our analytics pipeline and open-source software can be used to answer important prediction questions while limiting potential causes of bias (e.g., by validating phenotypes, specifying the target population, performing large-scale external validation and publicly providing all analytical source code).

**Methods:** We show step-by-step how to implement the pipeline for the question: ‘In patients hospitalized with COVID-19, what is the risk of death 0 to 30 days after hospitalization’. We develop models using six different machine learning methods in a US claims database containing over 20,000 COVID-19 hospitalizations and externally validate the models using data containing over 45,000 COVID-19 hospitalizations from South Korea, Spain, and the US.

**Results:** Our open-source tools enabled us to efficiently go end-to-end from problem design to reliable model development and evaluation. When predicting death in patients hospitalized for COVID-19 adaBoost, random forest, gradient boosting machine, and decision tree yielded similar or lower internal and external validation discrimination performance compared to L1-regularized logistic regression, whereas the MLP neural network consistently resulted in lower discrimination. L1-regularized logistic regression models were well calibrated.

**Conclusion:** Our results show that following the OHDSI analytics pipeline for patient-level prediction can enable the rapid development towards reliable prediction models. The OHDSI tools and pipeline are open source and available to researchers around the world.

## I. Introduction

THE COVID-19 pandemic continues to cause unprecedented pressure on healthcare systems worldwide, and many casualties at a global scale [1]. Due to the urgency of the COVID-19 pandemic there was increased pressure to efficiently develop COVID-19 prediction models. Unfortunately, model reliability was often compromised in order to rapidly develop models. Despite guidelines on developing and reporting of prediction models [2], there are several common problems identified in published COVID-19 prediction models including uncertain data quality, unclear target setting, lack of large-scale external validation, and insufficient reporting [3].

This motivates the need for a standardized approach for rapid yet reliable development and validation of prediction models, one that allows researchers to address various sources of bias. For instance, such an approach should ensure that the data used are representative of the population for which the developed prediction model is intended to be used in clinical practice, and that the quality of the phenotypes used is investigated and transparently reported.

Observational Health Data Sciences and Informatics (OHDSI) is an international, multi-stakeholder collaboration that has developed open-source solutions for large-scale analytics [4]. The OHDSI community has used these open-source solutions to generate observational evidence for COVID-19 and has impacted international clinical guidelines [5-16].

In this paper, we demonstrate the OHDSI analytics pipeline for patient-level prediction (henceforth referred to as “prediction pipeline”) as a standardized approach for reliable and rapid development and validation of prediction models. We show that our pipeline makes it possible to develop prediction models rapidly without compromising model reliability. The main contributions of our work are summarized as follows:

### 1. Reliable and rapid research

OHDSI implements a distributed data network strategy where no patient-level data are shared. Instead, the analytical source code is shared publicly, run by data partners on their data, and only aggregated results are shared with the study coordinator. Such a strategy has proven to enable international collaborative research while providing various advantages [17]. Key advantages of OHDSI’s distributed data network strategy that are particularly relevant for the current pandemic are:

- **Reliable research**. The use of open-source software tools and publicly shared analytical source code, along with extensive documentation makes studies conducted with the same analysis within this distributed data network fully *reproducible* (i.e., same data, same results), as well as *replicable* (i.e., similar data, similar results).
- **Rapid research**. To improve the interoperability of originally heterogenous observational data sources (e.g., electronic healthcare records (EHRs), administrative claims), they are mapped to a common data model (CDM). The use of an established CDM enables standardized approaches for data curation and enables standardized analytics pipelines to generate results much faster [9, 18, 19]. In addition, the data standardization enables the ability to externally validate models at scale to investigate how reliable the models are across different case-mixes

### 2. Analysis of COVID-19 data from multiple countries from around the world

In March 2020, the OHDSI community began contributing to generating observational evidence for COVID-19 with data from around the world. By November 2020, there were 22 databases (including EHRs, administrative claims, primary and secondary care databases) in the OHDSI network that incorporated COVID-19 data – 11 from North America, 8 from Europe, and 3 representing Asia-Pacific (Fig. 1). In total, the mapped data included:

**Fig. 1.**
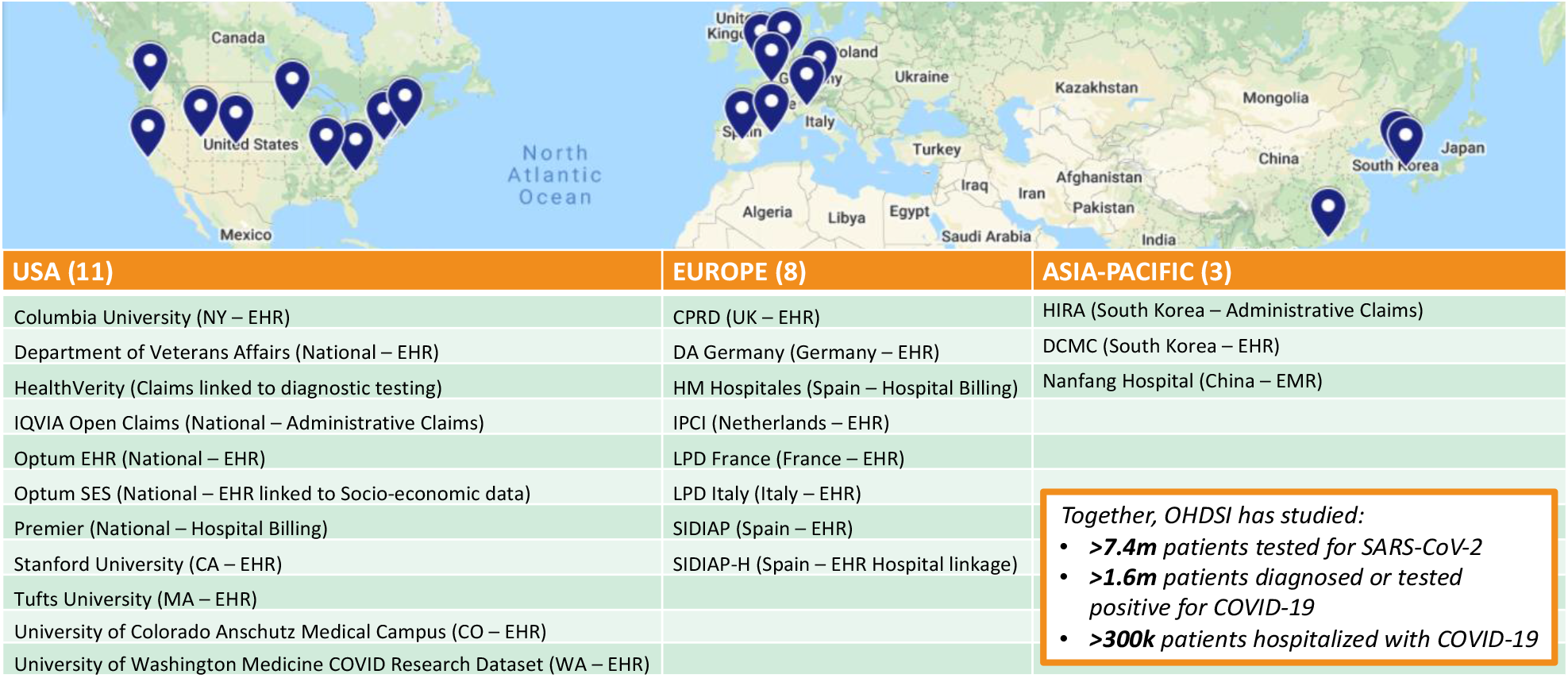
The OHDSI distributed data network. As of November 2020, it includes 22 sites spread across North America, Europe, and Asia that have COVID-19 patient data mapped to the Observational Medical Outcomes Partnership Common Data Model (OMOP CDM).

- > 7.4 million patients tested for severe acute respiratory syndrome coronavirus 2 (SARS-CoV-2)
- > 1.6 million patients diagnosed or tested positive for COVID-19
- > 300,000 patients hospitalized with COVID-19.

We describe each stage of the prediction pipeline in the following section. We then demonstrate the use of the prediction pipeline for the problem of predicting COVID-19 death in section III. This work was reviewed by the New England Institutional Review Board (IRB) and was determined to be exempt from board IRB approval, as this research project did not involve human subject research.

## II. METHODS

OHDSI provides a library of open-source software tools for the process of developing and validating prediction models using observational data (Fig. 2), beginning with data harmonization and quality control checks which are required before any database is added to the distributed data network and included in a study.

**Fig. 2.**
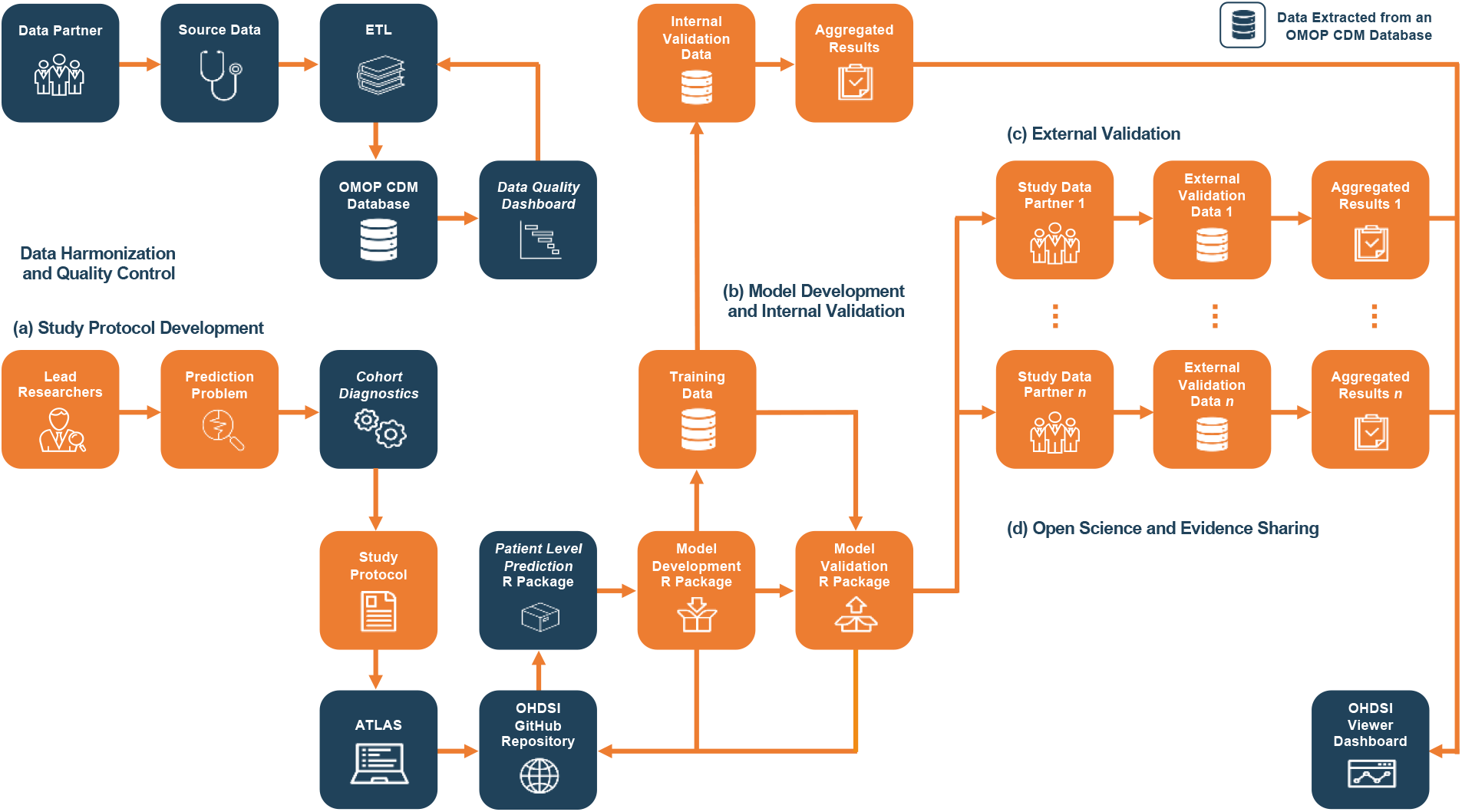
The OHDSI analytics pipeline for patient-level prediction modelling. As an initial check before a database is added to the distributed data network and included in a study, source data are harmonized to the OMOP CDM structure and coding system using an ETL design specification and quality control is performed. To conduct a prediction study, (a) we first develop a study protocol by defining the prediction problem, assessing phenotypes using the OHDSI CohortDiagnostics tool, and specifying machine learning settings; then (b) we develop and internally validate the prediction models using the ‘Model Development’ R package, a wrapper of the OHDSI PatientLevelPrediction R package, that we generate via the user-friendly interface in ATLAS; after which (c) we distribute the automatically generated ‘Model Validation’ R package to participating data partners for external validation of the developed models; finally (d) we disseminate our collected results on the OHDSI Viewer Dashboard. All documentation including the study protocol, generated R packages, and OHDSI tools, are publicly available on the OHDSI GitHub. Orange: study-specific input or output, blue: non-study-specific input, output, or OHDSI tool.

### Data Harmonization and Quality Control

OHDSI uses the Observational Medical Outcomes Partnership (OMOP) CDM which transforms source data into a common format using a set of common terminologies, vocabularies, and coding schemes [20]. To support the Extraction, Transformation and Load (ETL) of the source data to the CDM, the OHDSI community has developed several tools (Fig. 3).

**Fig. 3.**
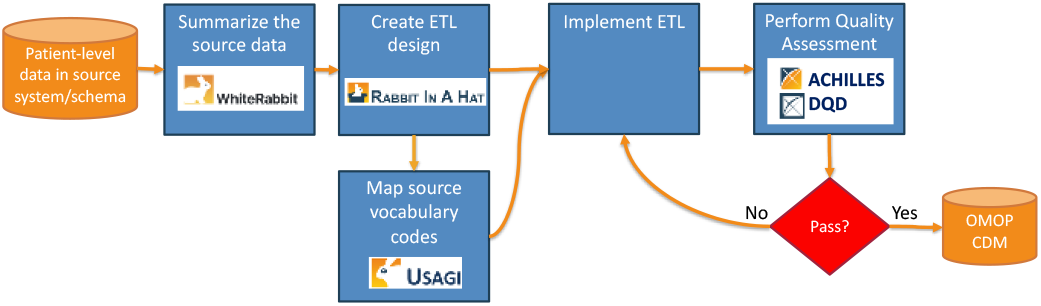
The step-by-step process for mapping data sources to the Observational Medical Outcomes Partnership Common Data Model (OMOP CDM), using OHDSI tools. ETL: Extraction, Transformation and Load; DQD: Data Quality Dashboard.

First, the WhiteRabbit tool produces a scan that summarizes every table, column, and value in a given source dataset [21]. This profiling step is important to understand the complexity of the source data.

Second, the Rabbit-In-A-Hat tool is an application for interactive design of an ETL to the OMOP CDM [22]. It reads the WhiteRabbit scan report and displays a graphical user interface containing all the source and OMOP CDM tables which need to be connected by the user. The final product is a design specification document that is then used to guide the implementation.

Third, the Usagi tool supports the mapping of source vocabularies to the OMOP standardized vocabularies [23]. Based on the ETL design specification that is created in Rabbit-in-a-hat the code is written to transform the source data into the OMOP CDM format.

Finally, the Data Quality Dashboard (DQD) and the ACHILLES characterization tool are used to interrogate the quality of the resulting OMOP CDM-mapped dataset [24]. The DQD uses a systematic approach to run and evaluate over 3,300 data quality checks. It assesses how well a dataset conforms with OMOP standards and how well source concepts are mapped to standard concepts. ACHILLES computes over 170 visualizations of the data and displays them in an open-source application designed to allow exploration and identification of potential anomalies and data outliers [25].

## Stages of the Study

### a) Study Protocol Development

Any research team from around the world can propose a study on the OHDSI Forum (https://forums.ohdsi.org). Interested investigators co-design a study protocol. The study protocol must transparently specify the prediction problem of interest and study design choices such as sensitivity analysis, model development methods, and evaluation techniques. Next, the collaborators determine the feasibility of the study across the data network and the validity of the specified study design choices using various OHDSI tools.

#### 1. Specifying the prediction problem

The OHDSI community has standardized the prediction problem specification into three components [26], which are shown in Fig. 4:

**Fig. 4.**
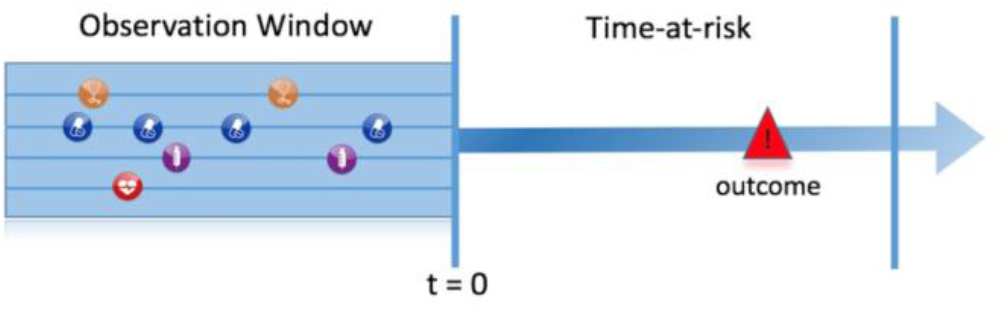
Prediction problem specification in OHDSI.

**Table I.** Prediction problem specification.

| Component | Specification |
| --- | --- |
| Target | Patients hospitalized with COVID-19 |
| Outcome | Death |
| Time-at-risk | 0 days to 30 days after the hospital visit |

- The target population – this is the set of patients for whom we wish to predict the individual risk. The index date (t=0) is the reference point in time for each patient in the target population. Only information from a specified observation window preceding the index date is used for engineering the candidate predictors.
- The outcome – this is the medical condition or event we wish to predict.
- The time-at-risk – this is a time interval on or after the index date, within which we wish to predict the outcome occurrence.

For example, if we wanted to develop a model to predict death within 30 days in patients hospitalized with COVID-19, then a suitable target population could be patients with a hospital stay who have COVID-19. The index date would be the first day of the hospital stay. The outcome would be a patient’s death, and the time-at-risk would be the period between index and 30 days after index.

#### 2. Generating and assessing phenotypes

Improving the syntactic and semantic interoperability of the data through the CDM and the standardized vocabularies does not solve all interoperability issues. For instance, data may originate from different clinical settings and have different levels of granularity. As a consequence, identifying the target population and outcome in the data can still be a challenge, even in observational data that are mapped to the OMOP CDM.

Defining an algorithm to identify patients within a database who have a certain condition or medical event is known as ‘phenotyping’. In general, a phenotype can be defined as an index rule followed by inclusion/exclusion rules. The rules can use sets of OMOP-standardized concept IDs to identify certain conditions or events. For example, our target population phenotype could be defined as follows.

Patients with an inpatient visit (concept ID 9201 or 262) satisfying the following inclusion criteria:

- COVID-19 positive test (concept ID 37310282) OR COVID-19 diagnosis (concept ID 439676, 37311061, 4100065 or 37311060) during the visit,
- ≥ 365 days of prior observation.

The index date is the date of the qualifying inpatient visit.

When developing a prediction model, it is important that the target population and outcome phenotypes correctly identify the desired individuals. A suitable phenotype is one that is highly sensitive (the majority of the patients in the database with the condition or event are correctly identified) and has a high positive predictive value (the majority of the patients identified by the phenotype have the condition or event). Mis-specifying the target population and/or outcome phenotype definitions is likely to lead to poor performance when implementing the prediction model in clinical practice. A significant amount of work is required to develop suitable phenotypes and expert knowledge of a specific database is required to guide this process.

The OHDSI community has developed a process to develop and evaluate suitable phenotypes. This process starts with a literature review of existing phenotype definitions for the target population and outcome. Commonly used phenotypes identified by the literature review are then identified as candidate phenotypes. If no phenotypes exist in the published literature, then a clinician and data expert collaborate to propose new candidate phenotypes. The candidate phenotypes need to be assessed to determine whether they are capturing the correct patients.

The OHDSI CohortDiagnostics tool creates descriptive results for each candidate phenotype such as the characteristics of the patients identified by the phenotype, the validity of the concept ID sets, and the number of patients identified by the phenotype across calendar time [27]. This is repeated across the OHDSI network of databases. The results are then inspected by a panel of clinicians to compare the characteristics of the patients identified by the phenotype, the temporal trend of the phenotype and the number of patients identified by the phenotype across numerous databases. The specific aspects of a phenotype that are inspected are:

- Generalizability. Do the patients captured by the phenotype appear to represent the real-world patients with the medical condition? This requires inspecting the characteristics of the patients identified by the phenotype in view of the literature and expert consensus.
- Consistency across the network. Is the phenotype identifying patients consistently across the network or does it seem to fail for one or more database? This may indicate an issue with the transportability of the definition. Common issues include unsuitable data or incorrect concept ID sets.
- Correctness of concept ID sets. Do we have the correct concept IDs for identifying inclusion/exclusion criteria used by the phenotype? OHDSI uses string similarity and associations to identify potential missing concept IDs.

If issues are identified with a candidate phenotype then revisions to the phenotype are made and the process is repeated until no issues are observed.

#### 3. Assessing suitability of source databases

Once the phenotypes are defined and validated, the next step is identifying whether each OHDSI observational database is suitable for model development and/or validation. This involves qualitative and quantitative assessment. If issues are identified, then other databases should be considered instead.

Initial feasibility assessment: Consulting with a person who has expert knowledge of the database is important to determine any issues in the way the data are captured that may impact the phenotypes. For example, some databases lack older or younger patients or may not capture complete lab results or medication.

Databases that pass the initial feasibility assessment are then reviewed using the CohortDiagnostics package. The results can be inspected to identify datasets that satisfy:

- Adequate size. Is the number of patients identified by the target population phenotype in a given database sufficient for developing a prediction model? Our recent but as yet unpublished study on generating learning curves to empirically assess the sample size at which convergence towards maximum achievable performance starts shows that, typically, more than 1000 patients with the outcome are needed for model development [28]. For accurate model validation, at least 100 patients with the outcome is recommended [29].
- Continuous observation time. Are the patients identified by the target population phenotype in a given database observed long enough to have a sufficient lookback period to capture predictors, and enough follow-up time to cover the time-at-risk? The incidence rate should be inspected for sufficient outcomes during the time-at-risk.

If there is no suitable database across the network then it may be worth exploring alternative prediction specifications (e.g., use a proxy for the target population).

#### 4. Model Development Settings

The study protocol must specify the settings used for model development including [26]:

- The candidate predictors to be included in the model, e.g., drugs (at various ingredient and drug levels), diagnoses, procedures, measurements, as well as diagnostics and summary scores.
- The train/test split design – by default a 25% test set and 75% train set is used, where *k*-fold cross validation is applied on the train set to select optimal hyper-parameters [30].
- The set of classifiers to be used, including gradient boosting machine, random forest, regularized logistic regression, decision tree, AdaBoost, and multi-layer perceptron neural network.
- The hyper-parameter search per selected classifier – if using a grid search the user can specify the values to investigate [30].
- Sensitivity analysis options – whether to include patients lost to follow-up or patients who had the outcome prior to index.

### (b) Model Development and Internal Validation

All model development settings can be specified via a user-friendly interface in ATLAS [4]. This includes the prediction problem components (the target population and outcome phenotypes and the time-at-risk) in addition to the above model specific settings (e.g., candidate predictor settings and model development settings).

#### 1. Developing the ‘Model Development’ R package

Once the settings have been specified, ATLAS generates a study-specific open-source R package called the ‘Model Development’ R package. This is a wrapper of the PatientLevelPrediction R package [26] and can be run on any OMOP CDM database to develop and internally validate the models specified in the study protocol.

#### 2. Executing the ‘Model Development’ R package

The ‘Model Development’ R package can be implemented by providing the connection details to the CDM, the CDM database name, a database schema with read/write access that is used to create temporary tables, and the location where the log/data/model will be saved to. The output is a directory containing the extracted data, the developed models, all the settings required to replicate the study and summary information about the internal validation of the models. In addition, an R Shiny app is generated that displays the results interactively to the user.

#### (c) External Validation

After developing the models, the ‘Model Development’ R package can automatically generate a ‘Model Validation’ R package for externally validating the models. This package contains the data extraction source code for the various settings (phenotypes, time-at-risk, and predictors) and the developed models that needs to be validated. The ‘Model Validation’ R package is another wrapper of the PatientLevelPrediction R package that uses the stored settings to call functions to extract the data, apply the models and assess the performance using the standard evaluation metrics. Therefore, the automatically generated ‘Model Validation’ R package is able to fully replicate the data extraction process used to develop the models across any database mapped to the OMOP CDM and then applies and validates the models. Once installed the user just points the ‘Model Validation’ R package to their OMOP CDM data and the R package will execute the external validation. The output is a collection of .csv files containing evaluation metrics such as information on the discrimination and calibration of each model.

### (d) Open Science and Evidence Sharing

All study documentation, including the study protocol and automatically generated R packages, are shared publicly. The ‘Model Development’ and ‘Model Validation’ R packages can be uploaded to the ohdsi-studies GitHub (https://github.com/ohdsi-studies) to enable any researcher to run the model development and external validation analysis on their data mapped to the OMOP CDM. Results for each of the databases participating in the study can be combined in an R Shiny application and then uploaded to the publicly available OHDSI Viewer Dashboard.

The OHDSI tools involved in the prediction pipeline are regularly updated and revised versions are maintained on the GitHub. The OHDSI Forum is open for all to join, to contribute to the development and use of tools, and to co-create scientific questions.

## COVID-19 Demonstration

In this section, we demonstrate using the prediction pipeline to develop and validate a COVID-19 prediction model. We were interested in predicting a patient’s risk of death within 30 days from the point they are hospitalized with COVID-19. We demonstrate how this was done using the stages of the prediction pipeline described in Section II.

### (a) Study Protocol Development

#### 1. Specifying the prediction problem

We studied the following prediction problem: “Within patients hospitalized with COVID-19, predict the risk of death on the hospitalization date and up to 30 days after using data recorded up to 1 day prior to hospitalization”, defined in Table I.

#### 2. Generating and assessing phenotypes

The phenotypes used to identify ‘patients hospitalized with COVID-19’ and ‘death’ are defined in Table II. The phenotype for ‘death’ was defined as any record of death in the database. The phenotype for ‘patients hospitalized with COVID-19’ was previously developed in a large-scale COVID-19 characteristic study detailed in https://github.com/ohdsi-studies/Covid19CharacterizationCharybdis. The CohortDiagnostics results are available at https://data.ohdsi.org/Covid19CharacterizationCharybdis/ for the cohort ‘Persons hospitalized with a COVID-19 diagnosis record or a SARS-CoV-2 positive test with at least 365d prior observation’. This phenotype was investigated across 16 OMOP CDM databases to ensure transportability (Fig. 5).

**Table II.** Phenotype definitions

| Component | Phenotype definition |
| --- | --- |
| Target | Patients with an inpatient visit on or after December 2019 with a COVID-19 positive test or COVID-19 diagnosis within 21 days before the visit or during the visit and $\geq 365$ days prior observation. |
| Outcome | Death record in database |

**Table 3.**
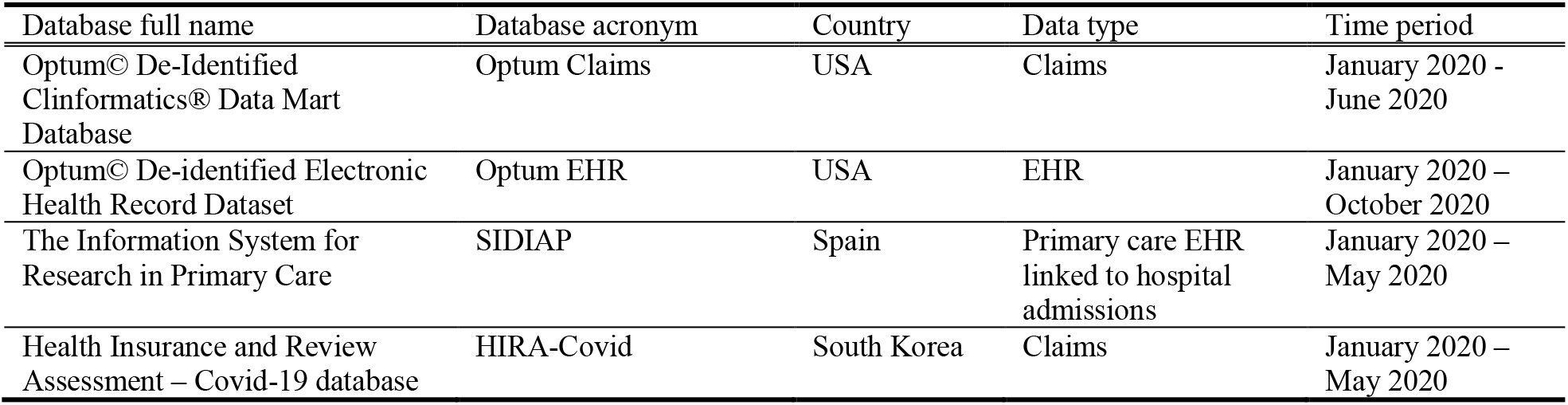
The databases used in this research.

| Database full name | Database acronym | Country | Data type | Time period |
| --- | --- | --- | --- | --- |
| Optum© De-Identified Clinformatics® Data Mart Database | Optum Claims | USA | Claims | January 2020 - June 2020 |
| Optum© De-identified Electronic Health Record Dataset | Optum EHR | USA | EHR | January 2020 – October 2020 |
| The Information System for Research in Primary Care | SIDIAP | Spain | Primary care EHR linked to hospital admissions | January 2020 – May 2020 |
| Health Insurance and Review Assessment – Covid-19 database | HIRA-Covid | South Korea | Claims | January 2020 – May 2020 |

**Table 4.**
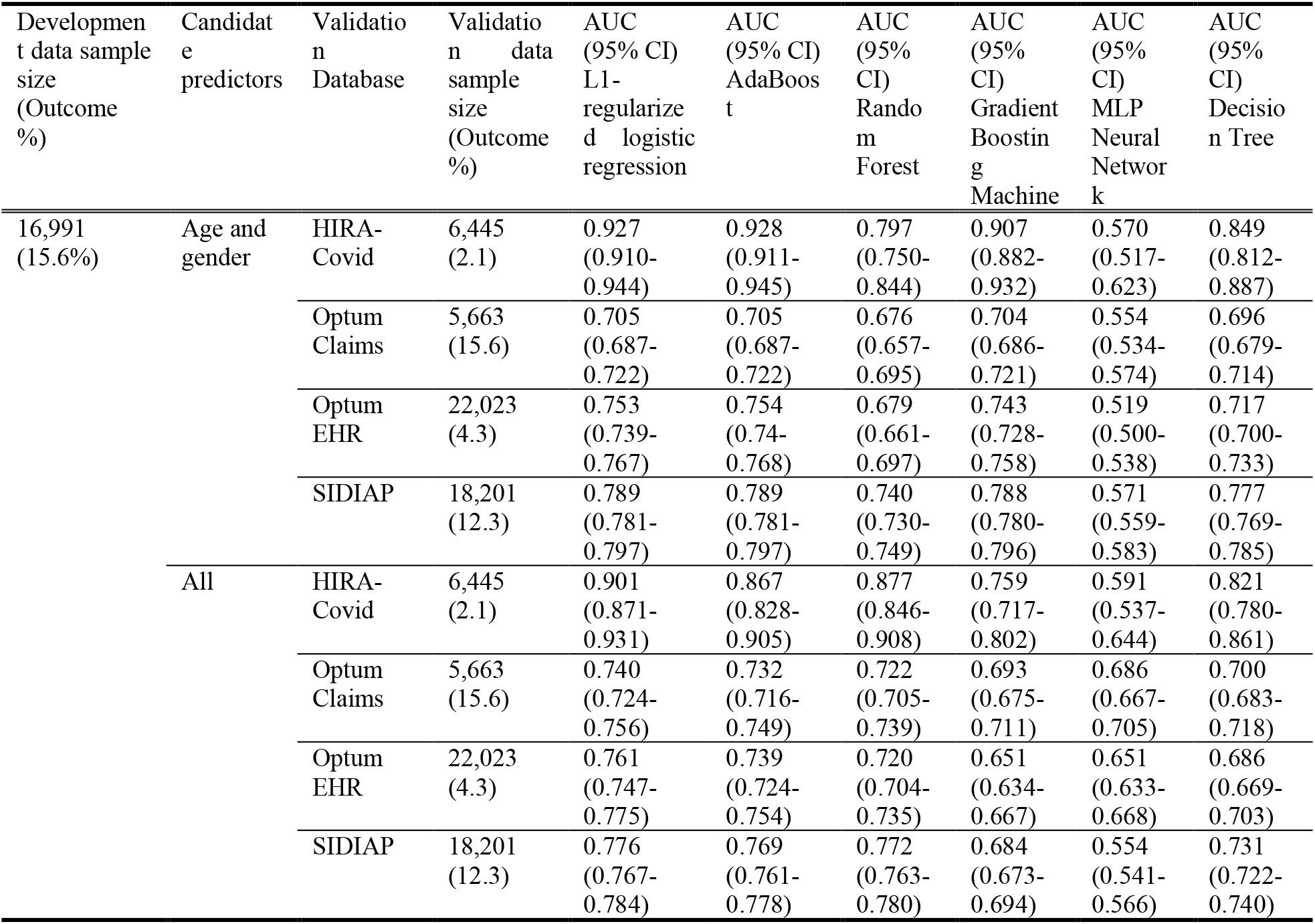
Discriminative performance (measured using the area under the receiver operating characteristic curve (AUC) with a 95% confidence interval (CI)) of the different classifiers in predicting 30-day death outcome in patients hospitalized with COVID-19 (Optum Claims is the internal discrimination estimated using the test set; the other databases are the external validation discrimination estimates).

#### 3. Assessing suitability of source databases

Across the OHDSI network, four OMOP CDM databases capturing death and containing inpatient visit data were identified as suitable, as per the database suitability checks (described in Section II) performed using the CohortDiagnostics tool. Table III describes the four databases. The largest one, Optum Claims was used to develop the models and Optum EHR, HIRA-Covid, and SIDIAP were used for external validation.

**Fig. 5.**
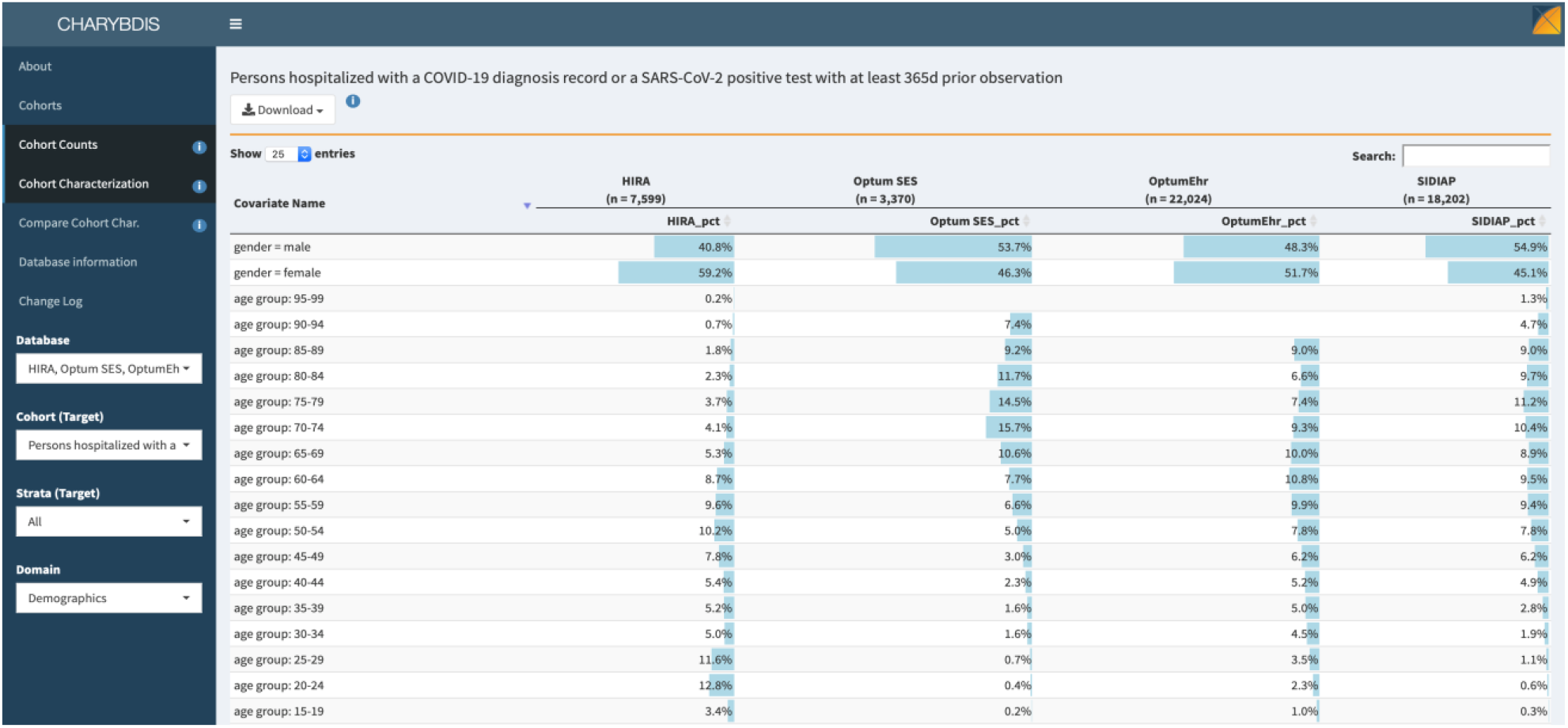
A snapshot of the CohortDiagnostics tool for assessing phenotypes. Here, Optum SES refers to the Optum Claims database.

#### 4. Model development settings

Two sets of candidate predictors were used.

- Age and gender: this set included gender and binary indicators of age in 5-year groups (40-45, 45-50, …, 95+). We used this set of candidate predictors to create a benchmark model.
- All: the second set included 57,627 candidate predictors including binary ones indicating the occurrences of various conditions, drugs, observations procedures or measurements, that were recorded any time prior, as well as in the year prior, to the index visit (not including day of the visit date), in addition to age and gender.

We chose a 75/25 train/test split with 3-fold cross-validation on the train set to select the optimal hyper-parameter settings per classifier. We trained a L1-regularized logistic regression model as the reference model, using cross-validation to select the strength of regularization. As a sensitivity analysis, we also trained Gradient Boosting Machine (GBM), decision tree, random forest, multi-layer perceptron (MLP) neural network, and AdaBoost models.

### (b) Model Development and Internal Validation

We developed the ‘Model Development’ R package in ATLAS (http://atlas-covid19.ohdsi.org/#/prediction/39). Within ATLAS, the phenotype definitions specified in Table II were created using the “Cohort Definitions” tab (Fig. 6). Next, the model settings were created using the “Prediction” tab. Once the prediction study was designed, the ‘Model Development’ R package was automatically generated by clicking on “Download Study Package”. The ‘Model Development’ R package contains all the functionality to develop and internally validate the prediction model using OMOP CDM data.

**Fig. 6.**
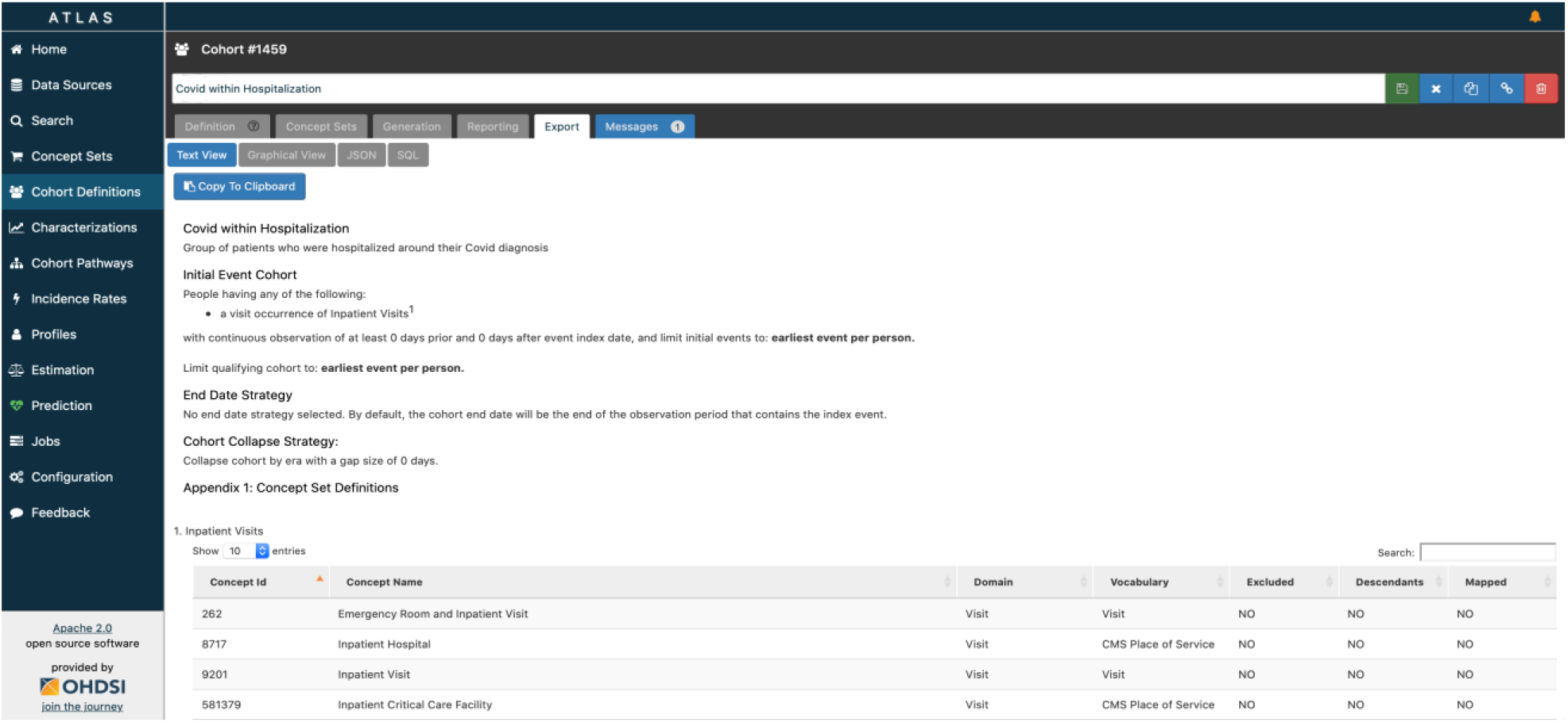
A snapshot of the ATLAS tool for prediction model development

### (c) External Validation

The ‘Model Validation’ R package was automatically generated using the ‘Model Development’ R package.

### (d) Open Science and Evidence Sharing

The protocol is available at: https://github.com/ohdsi-studies/CovidDeath/blob/master/inst/doc/protocol.docx.

The ‘Model Development’ R package is available at: https://github.com/ohdsi-studies/CovidDeath/tree/master/CovidDeathDev and the ‘Model Validation’ R package is available at https://github.com/ohdsi-studies/CovidDeath.

## III. RESULTS

Table IV presents the discriminative performance of the models. Using L1-regularized logistic regression, the model including the set of all variables resulted in an internal validation AUC of 0.74 (0.724-0.756) for Optum Claims and external validation AUCs of 0.761 (0.747-0.775) for Optum EHR, 0.776 (0.767-0.784) for SIDIAP, and 0.901 (0.871-0.931) for HIRA-Covid. In comparison, the L1-regularized logistic regression model including only age and gender predictors resulted in an internal validation AUC of 0.705 (0.687-0.722) for Optum Claims, and external validation AUCs of 0.753 (0.739-0.767) for Optum EHR, 0.789 (0.781-0.797) for SIDIAP, and 0.927 (0.910-0.944) for HIRA-Covid. For both sets of candidate predictors (age and gender only, and all variables), AdaBoost, random forest, gradient boosting machine, and decision tree yielded similar or lower internal and external validation AUCs compared to L1-regularized logistic regression, whereas the MLP neural network consistently resulted in lower AUCs. The models were well calibrated with respect to age and gender (Fig. 7 and Fig. 8).

**Fig. 7.**
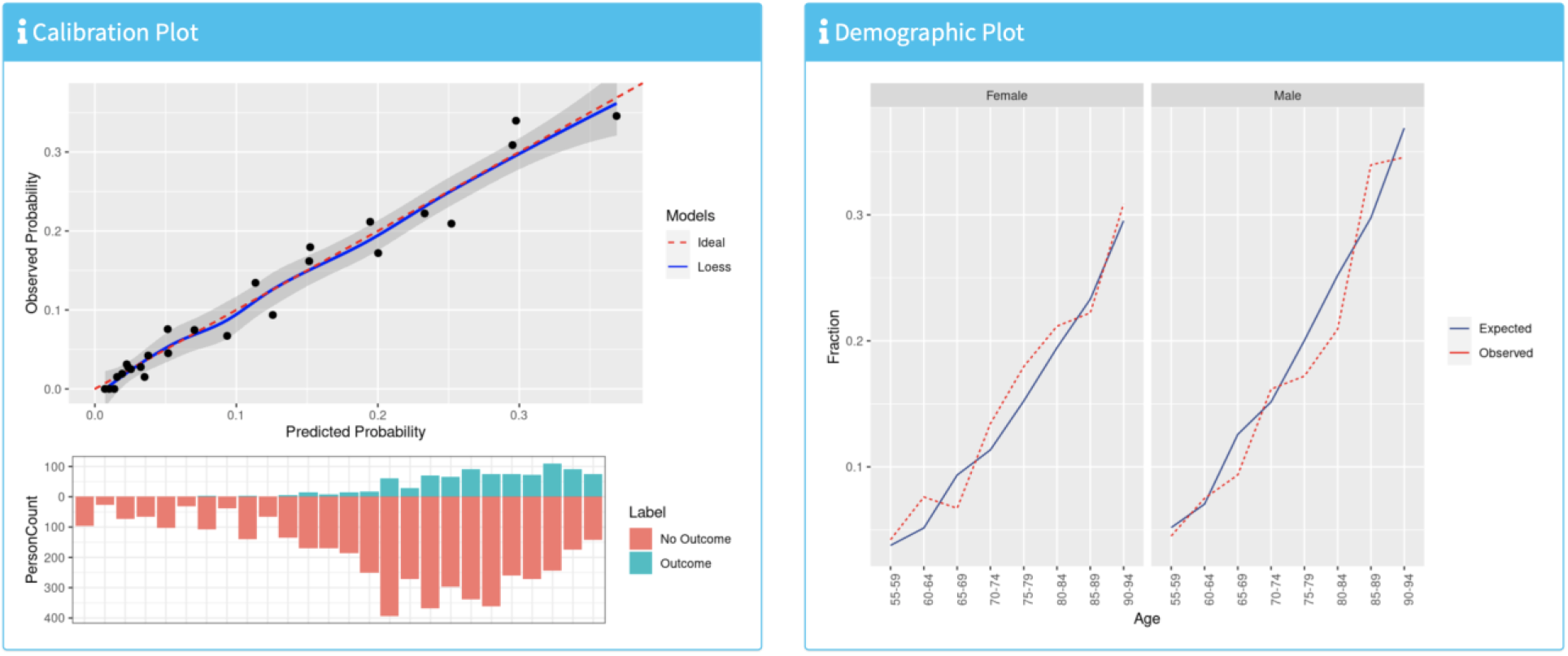
Calibration performance for internal validation of the L1-regularized logistic regression model for predicting 30-day death outcome in patients hospitalized with COVID-19 on Optum Claims data, overall (left panels) and by age and gender (right panels).

**Fig. 8.**
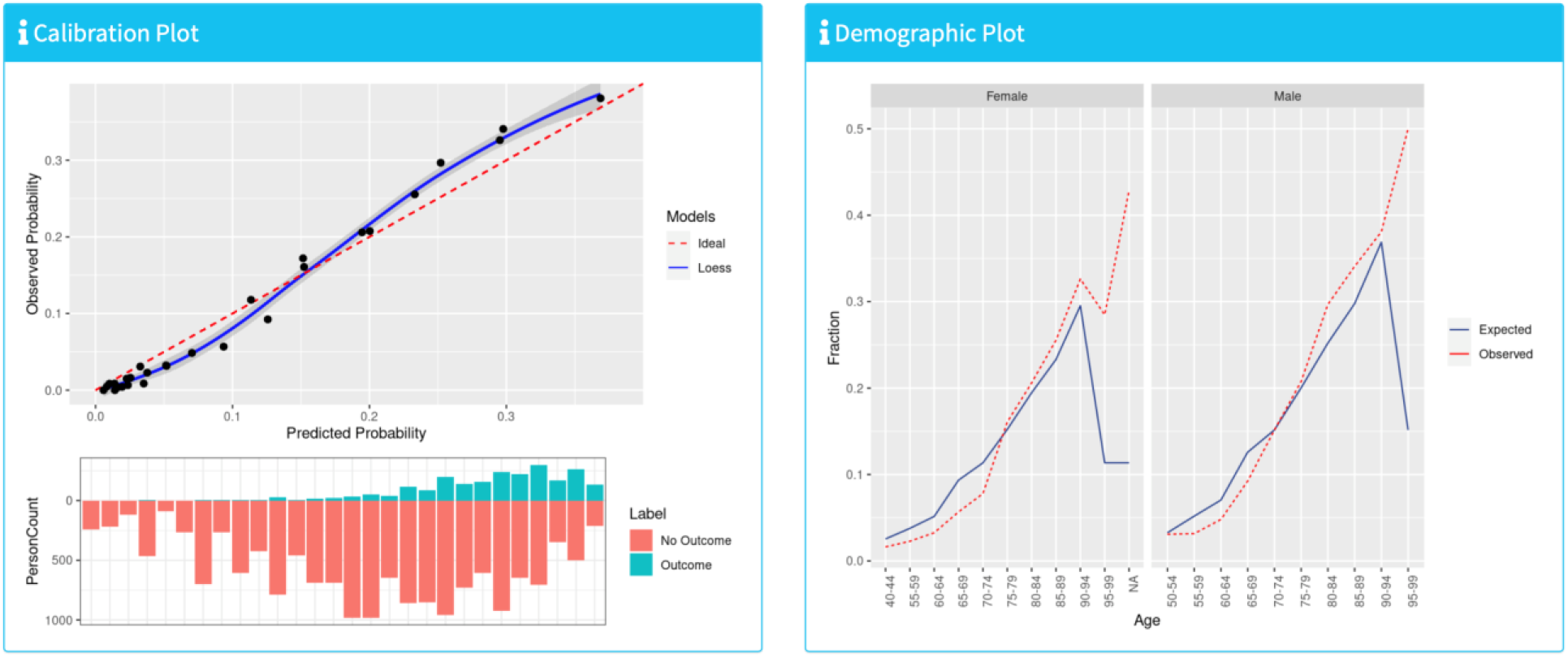
Calibration performance for external validation of the L1-regularized logistic regression model for predicting 30-day death outcome in patients hospitalized with COVID-19 on SIDIAP data, overall (left panels) and by age and gender (right panels).

The internal and external validation results were made publicly available in the OHDSI Viewer Dashboard at: https://data.ohdsi.org/CovidDeathPrediction/, which shows the *model summary* (including model coefficients or variable importance for non-generalized linear models), *model performance* (including discrimination (AUC, F1 Score, Precision (also known as Positive Predictive Value), Recall (also known as Sensitivity), and more) and calibration (observed vs predicted risk, overall and by age and gender)), and *all model settings* (Fig. 9). For instance, the hyperparameter values for all models used in this study are available in the “Settings” tab and Fig. 10 shows the intercept term and coefficients for the final age and gender L1-regularized logistic regression model in the “Model” tab. The complete model can also be downloaded from this tab.

**Fig. 9.**
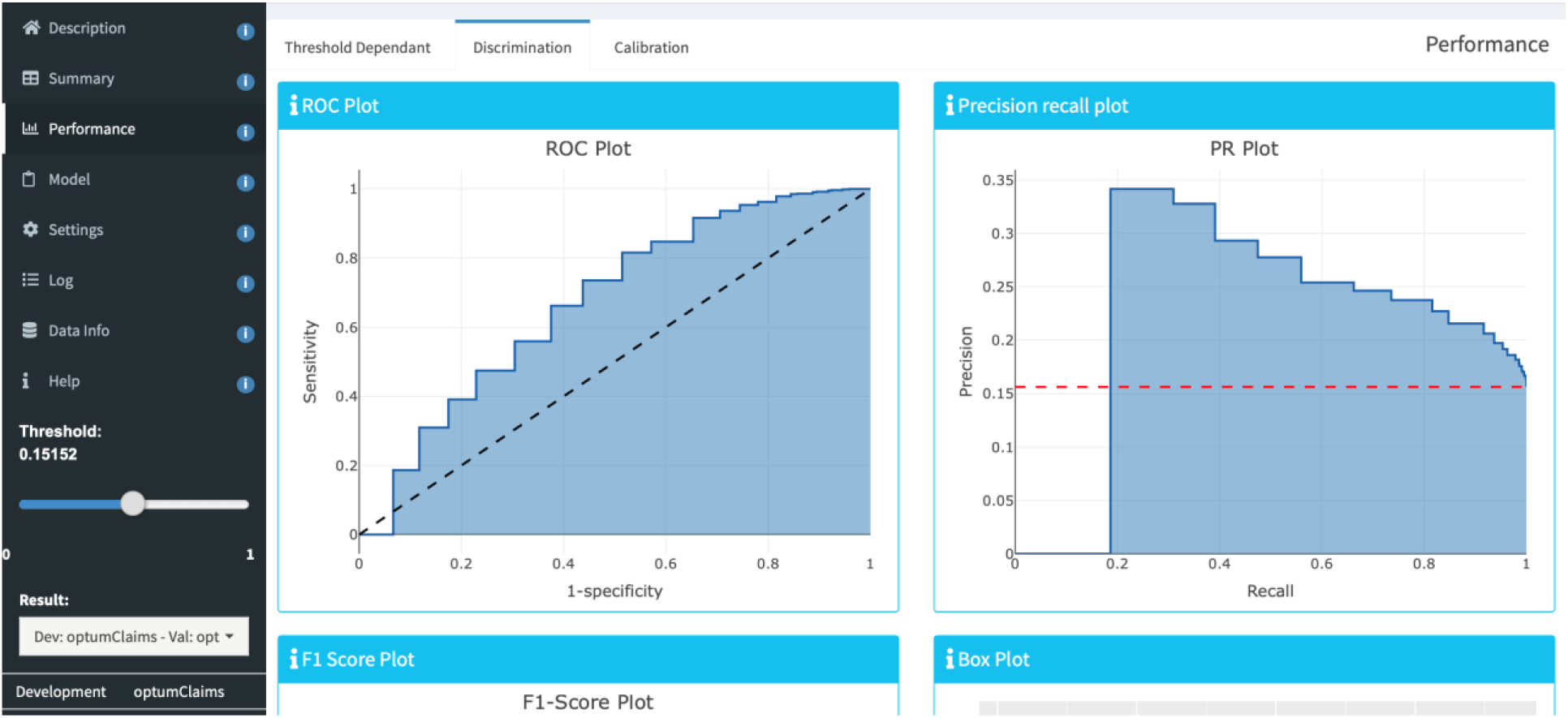
A snapshot of the Viewer Dashboard. It contains the model summary, model performance, and all model settings.

**Fig. 10.**
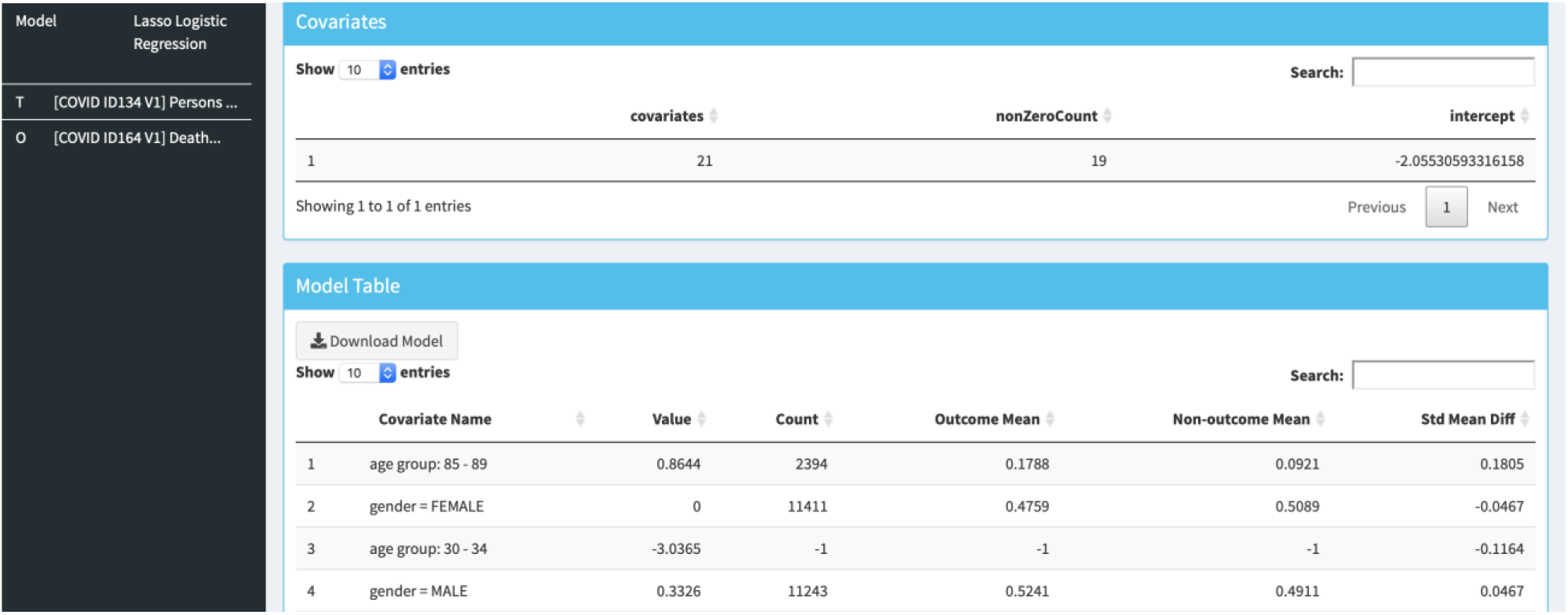
A snapshot of a Model Table in the Viewer Dashboard. It contains the complete model specification including intercept term and coefficient values for each covariate included in the final model.

## IV. Discussion

As an open science initiative, researchers anywhere in the world can join the OHDSI collaborative, and any data custodian can become a data partner by mapping their data to the OMOP CDM. The fast-growing OHDSI distributed data network enables performance assessment at a scale that can be highly valuable to develop prediction models that may impact patient care and outcome.

Once a prediction problem is specified, having suitable phenotypes and data is essential. Our prediction pipeline includes the important step of validating phenotypes across a network of databases prior to implementing any model development. This step aims to ensure phenotypes are transportable and aims to improve the reliability of the model. External validation across diverse datasets is made possible due to the OHDSI standardizations and collaborative network [31]. This is a key strength of our prediction pipeline and in this paper, we demonstrated how it was possible to perform external validation of prediction models across multiple countries. The majority of published COVID-19 prediction models were unable to provide such an extensive set of external validation results. Finally, our prediction pipeline enforces best practices for transparent reporting of prediction models as described in the TRIPOD statement [2].

Age and gender were found to be the main predictors of death within 30-days of hospitalization with COVID-19, which suggests our findings are consistent with literature [3]. Adding more variables improved the model performance in Optum Claims and Optum EHR, with the best performing model being L1-regularized logistic regression. Interestingly, the L1-regularized logistic regression, AdaBoost, and gradient boosting machine models that only used age and gender predictors performed the best in the SIDIAP data. In HIRA-Covid, we also found that the L1-regularized logistic regression models that only used age and gender predictors had the highest AUC of all models. This suggests that this model may be more transportable across countries and healthcare settings. However, although the Korean COVID-19 patient population itself is young, almost all deaths were in elderly patients over 65 years of age [5], and age being a dominant predictor is a possible reason for the better performance of the models using only age and gender. Further, a single measure cannot fully evaluate the model’s performance and other measures may provide a different interpretation. For instance, the AUPRC scores (in the Viewer Dashboard) for HIRA are lower than for the other databases, possibly due to a relatively low death rate in South Korea.

As with studies based on distributed data networks, a limitation of the approach presented in this paper is that it relies on data partners to map their data to the OMOP CDM. This initial mapping can be time-consuming. However, once done, a database can rapidly be integrated into a network study. Despite including data from three different continents, there are many regions of the world that are not represented in this paper. As more databases actively join the OHDSI data network (including from South Asia and Latin America), we can rapidly extend the external validation to them in the near future.

Using four databases from across the world, we developed and externally validated prediction models for 30-day risk of death in patients hospitalized with COVID-19. We demonstrated how the OHDSI analytics pipeline for patient-level prediction modelling offers a standardized approach for rapid yet reliable development and validation of prediction models, one that allows researchers to address various sources of bias. This work is a step towards obtaining prediction models that can provide reliable evidence-based guidance for use in clinical practice.

## Data Availability

All study documentation, including the study protocol and automatically generated R packages, are shared publicly. The Model Development and Model Validation R packages can be uploaded to the ohdsi-studies GitHub (https://github.com/ohdsi-studies) to enable any researcher to run the model development and external validation analysis on their data mapped to the OMOP CDM. Results for each of the databases participating in the study can be combined in an R Shiny application and then uploaded to the publicly available OHDSI Viewer Dashboard. The OHDSI tools involved in the prediction pipeline are regularly updated and revised versions are maintained on the GitHub. The OHDSI Forum is open for all to join, to contribute to the development and use of tools, and to co-create scientific questions.

## STATEMENTS OF ETHICAL APPROVAL

All databases obtained IRB approval or used deidentified data that was considered exempt from IRB approval. Informed consent was not necessary at any site.

